# Seroprevalence of anti-ganglionic acetylcholine receptor antibodies in patients with functional neurological symptom disorder/conversion disorder

**DOI:** 10.1101/2022.10.08.22280876

**Authors:** Ryusei Nagata, Eiji Matsuura, Satoshi Nozuma, Mika Dozono, Yutaka Noguchi, Masahiro Ando, Yu Hiramatsu, Daisuke Kodama, Masakazu Tanaka, Ryuji Kubota, Munekazu Yamakuchi, Yujiro Higuchi, Yusuke Sakiyama, Hitoshi Arata, Keiko Higashi, Teruto Hashiguchi, Shunya Nakane, Hiroshi Takashima

## Abstract

**Background:** Autoimmune autonomic ganglionopathy (AAG) is a rare disorder characterized by autonomic failure associated with the presence of anti-ganglionic acetylcholine receptor (gAChR) antibodies; however, several studies have reported that individuals with anti-gAChR antibodies present with central nervous system (CNS) symptoms such as impaired consciousness and seizures. In the present study, we investigated whether the presence of serum anti-gAChR antibodies correlated with autonomic symptoms in patients with functional neurological symptom disorder/conversion disorder (FNSD/CD).

**Methods:** Clinical data were collected for 59 patients presenting with neurologically unexplained motor and sensory symptoms at the Department of Neurology and Geriatrics between January 2013 and October 2017 and who were ultimately diagnosed with FNSD/CD according to the Diagnostic and Statistical Manual of Mental Disorders, 5th Edition. Correlations between serum anti-gAChR antibodies and clinical symptoms and laboratory data were analyzed. Data analysis was conducted in 2021.

**Results:** Of the 59 patients with FNSD/CD, 52 (88.1%) exhibited autonomic disturbances and 16 (27.1%) were positive for serum anti-gAChR antibodies. Cardiovascular autonomic dysfunction, including orthostatic hypotension, was significantly more prevalent (75.0% *vs* 34.9%, *p* = 0.008), whereas involuntary movements were significantly less prevalent (31.3% *vs* 69.8%, *p* = 0.007), among anti-gAChR antibody-positive compared with - negative patients. Anti-gAChR antibody serostatus did not correlate significantly with the frequency of other autonomic, sensory, or motor symptoms analyzed.

**Conclusions:** An autoimmune mechanism mediated by anti-gAChR antibodies may be involved in the etiology of FNSD/CD in a subgroup of patients.

## Introduction

Functional neurological symptom disorder/conversion disorder (FNSD/CD) is a neurological disorder of voluntary movement or sensory function; and is described in the Diagnostic and Statistical Manual of Mental Disorders, Fifth Edition (DSM-5); as a long-term impairment that significantly impairs quality of life and requires assistive and supportive services ^1^. Because the etiopathogenesis of FNSD/CD is unknown and abnormalities in clinical tests are rare, FNSD/CD is generally considered to be a psychological disorder ^2 3^. It is sometimes difficult to diagnose patients with nonspecific motor-sensory symptoms as FNSD/CD, and some patients initially diagnosed with FNSD/CD are eventually diagnosed with tumor-associated neurological syndrome or autoimmune encephalitis^3 4 5 6^. In addition, elevated inflammatory markers in the blood have been reported in some FNSD/CD patients ^7 8^, leading to the hypothesis that FNSD/CD symptoms may correlate with immune dysfunction. To avoid misdiagnosis, physicians are recommended to not assign a diagnosis of psychogenic reactions to neurological symptoms that are difficult to explain, such as dystonia or atypical seizures. Indeed, DSM-5 and International Classification of Diseases, 11th Revision (ICD-11) no longer use the term psychogenic factors as a cause of symptoms of neurological disorders such as involuntary movement.

While patients with FNSD/CD often exhibit autonomic symptoms such as orthostatic hypotension and gastrointestinal and dysuria, the exact frequency of autonomic complications in patients with FNSD/CD is unknown. ^9 10^. The autonomic nervous system is broadly divided into sympathetic and parasympathetic components, with the main neurotransmitters being norepinephrine and acetylcholine (ACh). The two ACh receptor (AChR) subtypes, nicotinic and muscarinic, are widely expressed throughout the body. In both the sympathetic and parasympathetic ganglia, preganglionic to postganglionic fiber neurotransmission is predominantly mediated by ACh and the major receptor involved is the nicotinic AChR (nAChR) ^11^. nAChRs exist as homodimeric or heteromeric complexes of nine subunits (α2–7 and β2–4) in peripheral nerves and the central nervous system of mammles ^12^. In humans, autonomic ganglia nAChRs generally exist as pentameric heterocomplexes, with α3 and β4 subunits playing important roles ^13 14^. Autoantibodies against nAChRs have been associated with several diseases, including myasthenia gravis, in which the antibodies target the α1 subunit of nAChRs at the neuromuscular junction ^15^. Serum antibodies against the autonomic ganglion nAChR (anti-gAChR) were first detected in patients with autonomic dysfunction in 1998 ^16^ and resulted in the concept of autoimmune autonomic ganglionopathy (AAG), a term that refers to autonomic neuropathy mediated by an autoimmune mechanism.

In the present study, we retrospectively reviewed clinicopathological data, including autonomic, motor, and sensory symptoms and anti-gAChR antibody serostatus, in patients who were admitted to our Neurology Department with neurologically unexplainable motor and sensory symptoms and were ultimately diagnosed with FNSD/CD.

## Methods

### Patients

Patients with the main complaint of motor and sensory disorders who were admitted to the Department of Neurology and Geriatrics at Kagoshima University Hospital between January 2013 and October 2017 were included in this study. Of those patients, 68 met the DSM-5 diagnostic criteria for FNSD/CD ^1^. The medical records of the subjects were retrospectively reviewed for clinical symptoms, blood and cerebrospinal fluid (CSF) test results. After exclusion of patients with concomitant autoimmune neurological disease, a total of 59 patients were included in the final analysis set. This study was approved by the Ethics Review Board on Bioethics and Genetic Research Reference of Kagoshima University (approval number 492). Written informed consent was obtained from the patients.

### Neurological examinations

Autonomic symptoms were categorized into four types: gastrointestinal dysfunction (constipation, diarrhea, early satiety, nausea, vomiting, anorexia, abdominal pain, and feeling abdominally bloated); cardiovascular autonomic dysfunction (orthostatic hypotension, postural orthostatic tachycardia syndrome [POTS], tachycardia, and palpitations); bladder dysfunction; and anhidrosis. Motor symptoms were categorized into three types: weakness, involuntary movements, and ataxia. Sensory symptoms were categorized as either sensory disturbance (abnormal or decreased sensation) or pain (headache, pain in extremities and trunk).

### Detection of anti-gAChR antibodies using a luciferase immunoprecipitation system (LIPS)

Serum antibodies against gAChR α3 and β4 subunits were quantified using a LIPS assay ^13 17^, in which specific antibodies are bound to a *Gaussia* luciferase-tagged antigen and the bound antibody is quantified by measuring luciferase activity with a luminometer. The results are expressed as relative luminescence units (RLU). Serum was considered positive for anti-gAChR α3 or β4 antibodies if the RLU of the test sample exceeded a cut-off value of three standard deviations (SDs) greater than the mean RLU of the control sample (sera obtained from healthy individuals).

### Statistical analysis

Mean values were compared between anti-gAChR antibody-positive and -negative patient groups. Age and laboratory findings were compared using *t*-tests, and gender, clinical symptoms, and other autoantibodies were compared using Fisher’s exact test. IBM SPSS, version 27, was used for statistical analyses. *p* < 0.05 was considered significant.

## Results

Of the 68 patients who presented with motor or sensory deficits and met the diagnostic criteria for FNSD/CD, 9 had concomitant neuroimmune disease and were excluded. The final analysis set consisted of 59 patients (Table 1), of whom 51 (86.4%) were female. The mean (± SD) age at admission was 26.4 ± 15.4 years, the age at symptom onset was 22.0 ± 14.9 years, and the duration of illness was 4.4 ± 5.9 years. We examined the prevalence of motor, sensory, and autonomic symptoms, and found that 45 of the 59 patients (76.3%) had all three symptom types, one (1.7%) had only motor symptoms, and none had only sensory symptoms (Fig 1). Of the 59 patients, 52 (88.1%) had autonomic symptoms that included gastrointestinal symptoms (40 patients, 67.8%), cardiovascular autonomic symptoms (27 patients, 45.8%), bladder dysfunction (23 patients, 39.0%), and anhidrosis (16 patients, 27.1%) (Table 1).

**Table 1.**
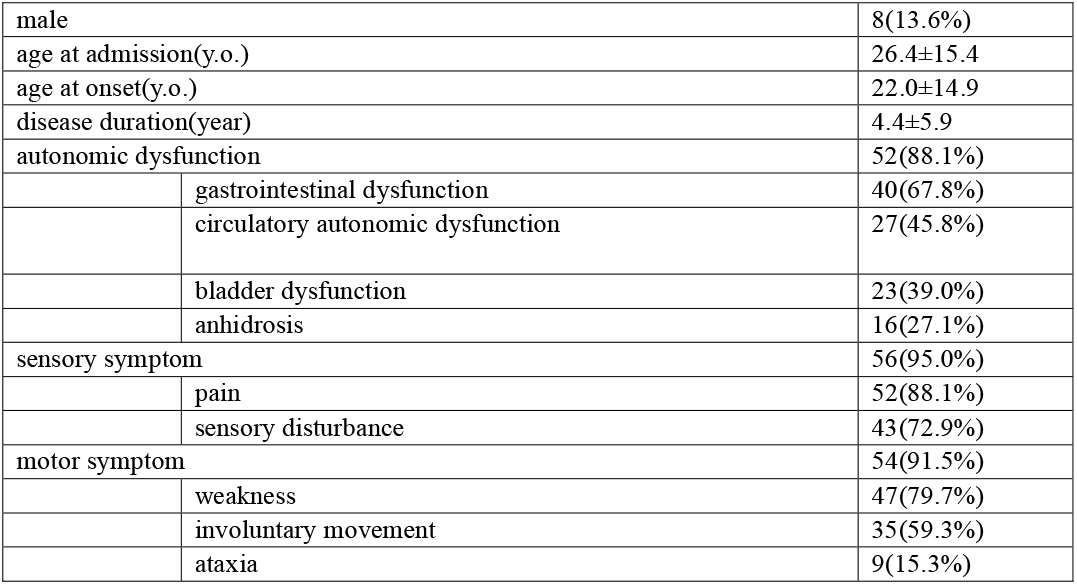
Characteristics of patients with FNSD/CD (*N* = 59)

**Figure 1.**
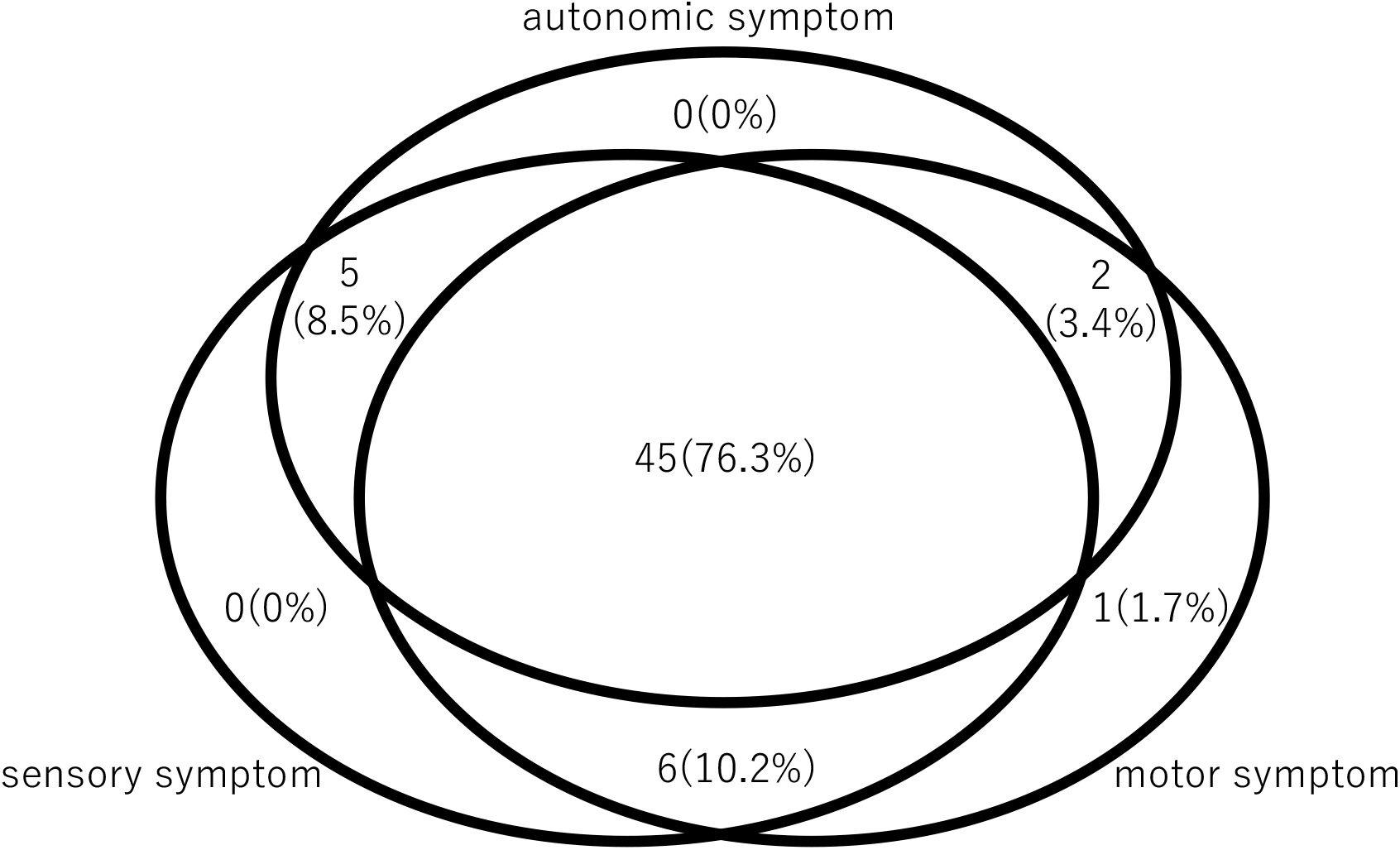
Symptoms of 59 patients with FNSD/CD. Prevalence of symptoms in 59 patients with FNSD/CD. Venn diagram shows the number and percentage of patients with autonomic, sensory, and motor symptoms.

Because of the prevalence of autonomic symptoms among our patient cohort, we investigated the possible relationship between FNSD/CD and anti-gAChR antibodies. Of the total cohort of 59 patients, 16 (27.1%) were positive for anti-gAChR antibodies, and of these, 11 were positive for antibodies against the α3 subunit only, one patient was positive for antibodies against the β4 subunit only, and four patients were positive for antibodies against both the α3 and β4 subunits (Fig 2).

**Figure 2.**
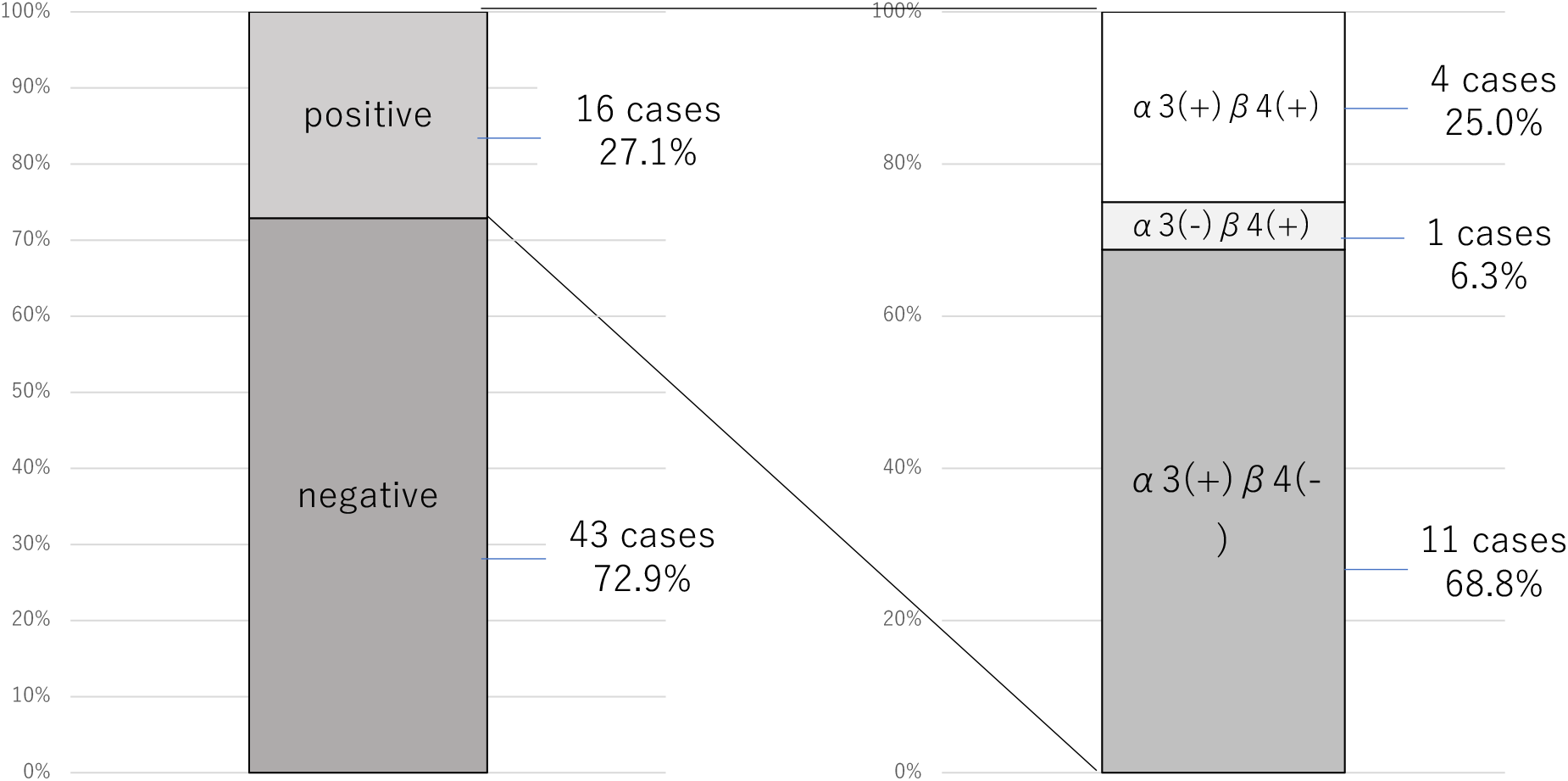
Anti-gAChR antibodies in patients with FNSD/CD. Prevalence of anti-gAChR antibodies in 59 patients with FNSD/CD. The proportion of patients positive for anti-gAChR antibodies (left) and for antibodies specifically against the α3 and β4 subunits (right) are shown.

We next compared clinical symptoms between patients who were positive (n=16) and negative (n=43) for anti-gAChR antibodies (Table 2). There were no significant differences between the antibody-positive and -negative groups with respect to sex ratio (3 males [18.8%] and 5 males [11.6%], respectively), mean age at admission (30.7 ± 17.8 years and 24.8 ± 14.5 years, respectively, or mean age at disease onset (23.3 ± 16.8 years and 21.5 ± 14.4 years, respectively). There was no significant difference in the prevalence of autonomic dysfunction between the anti-gAChR antibody-positive group (15 of 16 patients, 93.8%) and the antibody-negative group (37 of 43 patients, 86.0%); however cardiovascular autonomic symptoms were significantly more common in the antibody-positive group (75.0% *vs* 34.9%, *p* = 0.008). Among the individual cardiovascular autonomic symptoms, POTS and orthostatic hypotension were both more commonly seen among the antibody-positive group, but only the frequency of POTS reached the level of statistical significance (50.0% *vs* 9.3%, *p* = 0.002). There was no significant association between anti-gAChR antibody serostatus and other autonomic dysfunctions (Table 2).

**Table 2.** Characteristics of FNSD/CD patients according to anti-gAChR antibody serostatus

|  |  | anti-gAChR antibody |  |  |
| --- | --- | --- | --- | --- |
|  |  | positive(N=16) | negative(N=43) | p value |
| male |  | 3(18.8%) | 5(11.6%) | 0.670 |
| age at admission(y.o.) |  | 30.7±17.8 | 24.8±14.5 | 0.192 |
| age at onset(y.o.) |  | 23.3±16.8 | 21.5±14.4 | 0.699 |
| disease duration(year) |  | 7.5±8.8 | 3.2±4.0 | 0.079 |
| autonomic dysfunction |  | 15(93.8%) | 37(86.0%) | 0.661 |
| gastrointestinal dysfunction |  | 12(75.0%) | 28(65.1%) | 0.545 |
| cardiovascular dysfunction |  | 12(75.0%) | 15(34.9%) | 0.008 |
| orthostatic hypotension |  | 8(50%) | 9(20.9%) | 0.05 |
| POTS |  | 8(50%) | 4(9.3%) | 0.002 |
| palpitation |  | 2(12.5%) | 3(7.0%) | 0.606 |
| bladder dysfunction |  | 7(43.8%) | 16(37.2%) | 0.647 |
| anhidrosis |  | 7(43.8%) | 9(20.9%) | 0.104 |
| sensory symptom |  | 14(87.5%) | 42(97.7%) | 0.176 |
| pain |  | 13(81.3%) | 39(90.7%) | 0.375 |
| sensory disturbance |  | 12(75.0%) | 31(72.1%) | 1.000 |
| motor symptom |  | 14(87.5%) | 40(93.0%) | 0.606 |
| weakness |  | 12(75.0%) | 35(81.4%) | 0.718 |
| involuntary movement |  | 5(31.3%) | 30(69.8%) | 0.007 |
| ataxia |  | 2(12.5%) | 7(16.3%) | 1.000 |

POTS: postural orthostatic tachycardia syndrome

We next compared the motor and sensory symptoms between the anti-gAChR antibody-positive and -negative groups. Notably, although most motor and sensory symptoms were less common in the anti-gAChR antibody-positive group than the antibody-negative group, only the frequency of patients with involuntary movements was significantly different between the two groups (31.3% *vs* 69.8%, *p* = 0.007) (Table 2). The results of common laboratory tests revealed no significant differences in peripheral blood inflammatory markers, such as white blood cell count and C-reactive protein level, or in CSF inflammatory markers, such as total cell count and total protein level, between the anti-gAChR antibody-positive and -negative patient groups. The IgG index, which measures the ratio of IgG and albumin levels in the CSF *vs* serum and is an indicator of IgG production in the CNS, was also within normal limits in both patient groups (Table 3). Analysis of serum autoantibodies revealed that the presence of anti-nuclear antibodies was significantly more common in the anti-gAChR antibody-positive than -negative group (46.7% *vs* 12.5%, *p* = 0.006).

**Table 3.** Laboratory data for FNSD/CD patients according to anti-gAChR antibody serostatus

|  | anti gAChR antibody |  | p value |
| --- | --- | --- | --- |
|  | positive | negative |  |
| serum WBC count(/μl) | 5800.0±1065.6 | 6248.1±1645.9 | 0.317 |
| serum CRP(mg/dl) | 0.101±0.178 | 0.037±0.447 | 0.147 |
| CSF cell count(/μl) | 1.25±0.45 | 1.22±0.47 | 0.826 |
| CSF protein(mg/dl) | 22.60±6.45 | 24.40±11.97 | 0.573 |
| IgG index | 0.490±0.041 | 0.483±0.046 | 0.597 |
| anti nucler antibody | 7/15(46.7%) | 5/40(12.5%) | 0.006 |
| anti Tg antibody | 0/15(0%) | 3/40(7.5%) | NA |
| anti TPO antibody | 0/15(0%) | 3/41(7.3%) | NA |
| anti GAD antibody | 0/7(0%) | 0/25(0%) | NA |
| anti SS-A antibody | 1/12(8.3%) | 0/39(0%) | NA |
| anti SS-B antibody | 0/13(0%) | 0/38(0%) | NA |

Tg: thyroglobulin; TPO: thyroid peroxidase; GAD: glutamic acid decarboxylase

## Discussion

Among our cohort of FNSD/CD patients, more than two-thirds (88.1%) presented with autonomic symptoms and, notably, more than a quarter (27.1%) were positive for serum anti-gAChR antibodies. Autonomic symptoms have been reported to be a precursor to FNSD/CD and have been positively associated with autism ^18 10^. However, to our knowledge, there have been no formal investigations of autonomic dysfunction in patients with FNSD/CD, possibly because the DSM-5 diagnostic criteria for FNSD/CD require the presence of motor and sensory symptoms but not of autonomic symptoms. Although motor and sensory symptoms in FNSD/CD patients can be ruled in or out as consistent with neurological dysfunction by neurologists, it is more difficult to rule out a neurological basis for autonomic symptoms. In the present study, we found high rates of autonomic symptoms (gastrointestinal and cardiovascular neuropathy) in addition to motor and sensory symptoms, were the main symptoms of FNSD/CD, suggesting that evaluation of autonomic symptoms is important for both the diagnosis of FNSD/CD and for understanding its pathophysiology.

Anti-gAChR antibodies were initially found in the serum of patients with autonomic symptoms, leading to the discovery of autoimmune autonomic neuropathy, however, these antibodies are usually rarely detected. McKeon et al. reported that serum anti-gAChR antibodies were present in 155 of 15,000 patients (1.03%) with suspected paraneoplastic autoimmune neurological syndrome but in only 1 of 173 healthy subjects (0.58%) ^19^. More recently, 0% of 2628 patients with non-AAG neurological diseases were positive for antibodies to α3-nAChR ^20^. These reports suggest that anti-gAChR antibodies are usually barely detectable in the serum. ^16 17 21^. The high prevalence of anti-gAChR antibodies among our FNSD/CD patient cohort (27.1%) prompted us to hypothesize that anti-gAChR antibodies may be directly associated with autonomic symptoms in these patients. However, we detected no significant difference in the frequency of autonomic symptoms between patients who were positive or negative for serum anti-gAChR antibodies, suggesting that a direct relationship between the antibodies and autonomic symptoms is unlikely.

Anti-nuclear antibodies were significantly elevated in patients positive for anti-gAChR antibodies in the present study. In the past, some patients had antibodies against nAChR α4 and α7 subunits, VGKC channels, amphiphysin, and SS-B detected simultaneously with anti-gAChR antibodies, suggesting that a variety of humoral immunity may be activated in the patient’s serum ^27 28 29^. Interestingly, some patients with CNS symptoms such as impaired consciousness and seizures are positive for anti-gAChR antibodies, and several reports suggest that α7 and α4β2 nAChRs are involved in memory and pain^12 16 19 22 23 24 25 26^. Notably, recent fMRI studies have revealed that disturbances in the central autonomic network (CAN) can lead to psychiatric symptoms^30 31 32^. These facts suggest that antibodies targeting nAChR subunits can affect not only the peripheral autonomic nervous system but also CNS function. The detection of anti-gAChR antibodies in patients with FNSD/CD with or without autonomic symptoms in the present study suggests that anti-gAChR antibodies do not simply affect peripheral autonomic function.

## Conclusions

More than 80% of our patient cohort had autonomic symptoms, and anti-gAChR antibodies were detected in more than 25% of patients with FNSD/CD, whether or not autonomic symptoms were present. To clarify the relationship between this psychiatric disorder and anti-gAChR antibodies, further investigation of clinical symptoms and the establishment of a measurement system for antibodies against the anti-nAChR subunit are needed.

## Data Availability

All data produced in the present study are available upon reasonable request to the authors

## Acknowledgments

The authors thank the staff of the Department of Neurology and Geriatrics, Kagoshima University Hospital, including Katsunori Takahata, Yoshimitsu Maki, Michiyoshi Yoshimura, Yuichi Tashiro, Akihiro Hashiguchi, and Kumiko Michizono. We thank Anne M. O’Rourke, PhD, from Edanz (www.edanz.com/ac) for editing a draft of this manuscript.

## Author contributions

RN, EM, and HT conceived and designed the study. SNo, MD, YN, MA, YHir, YHig, YS, HA and KH provided the laboratory data for patients at Kagoshima University Hospital. SNa provided the laboratory data for control subjects. KD, MT, RK, MY and TH analyzed the data and take responsibility for the integrity of the data and the accuracy of the data analysis. All authors critically revised the manuscript and approved of the final version.

